# Effect of rasagiline on balance in Parkinson’s disease as measured by computerized posturography

**DOI:** 10.1101/2025.07.25.25331320

**Authors:** Fekete Robert

**Affiliations:** Department of Neurology, New York Medical College, Valhalla, NY, United States of America

**Author notes:** Corresponding Author Dr. Robert Fekete, 19 Bradhurst Ave, Hawthorne, NY 10532, United States of America.

**Keywords:** rasagiline, balance, posturography, Parkinson’s disease

## Abstract

**Background:** Falls occur in a majority of Parkinson’s disease (PD) patients. Balance disturbance can be measured by computer dynamic posturography. The effect of monoamine oxidase inhibition on PD patients as measured by computer dynamic posturography has not been previously studies.

**Methods:** The study will test the hypothesis that therapy with rasagiline improves balance in PD patients as measured by computerized dynamic posturography. 5 subjects will be randomized to rasagiline or placebo and 5 further subjects will be randomized to rasagiline as adjuvant therapy versus placebo. Subjects will be analyzed by computerized dynamic posturography testing. The device measures body sway under varying visual and vestibular cues and is used for assessment of balance. Subjects will be evaluated at baseline prior to taking first tablet as well as at 4 weeks and 8 weeks. In the adjuvant therapy arm, subjects will be evaluated on rasagiline but in a functional OFF state for their other PD medications. If the scores are normally distributed, we will determine whether data variance is approximately equal in all tested groups. If these conditions are met, data evaluation will continue using repeated measures ANOVA with between factor of drug (levels: rasagiline, placebo) and within factor of treatment (levels: before, after). Level of significance will be preset to 0.05.

## Background

Posturography testing for Parkinson’s disease patients on monoamine oxidase (MAO) inhibitors in general has not been studied previously. In Wood’s study of patients with Parkinson’s disease, falls occurred in 68.3% of the subjects [1]. In Ebmeier’s population study of idiopathic Parkinson’s disease patients, signs of postural instability on clinical examination were among the factors listed in premature death [2].

## Hypothesis

The hypothesis is that therapy with rasagiline improves balance in PD patients as measured by computerized dynamic posturography.

## Methods

Clinical trial number: not applicable. The study was not registered as no patients were enrolled. The study was approved by the New York Medical College Office of Research Administration Institutional Review Board under tracking number L-10,768.

## Inclusion Criteria

1. 18 years old or above
2. Clinical diagnosis of Parkinson disease by verified by movement disorders expert at the initial study visit with at least two cardinal signs of the disease (rest tremor, bradykinesia, rigidity, and postural instability).
3. For the monotherapy arm, patients must not be on amantadine, dopamine agonists, or levodopa. For the adjuvant therapy arm: Patients must be on a stable dose of their current medication for treatment of Parkinson disease which may include any combination of the following: amantadine, trihexiphenydil, dopamine agonist, and/or levodopa.
4. Patients may continue their stable dose of tricyclic, selective serotonin reuptake inhibitor, or serotonin norepinephrine reuptake inhibitor if they are on these medications at randomization.

## Exclusion Criteria

1. Catechol-O-Methyltransferase (COMT) inhibitor therapy use 30 days prior to start of study (both study arms).
2. Dopamine receptor blocker use (such as quetiapine) one week prior to taking study drug
3. For both monotherapy and adjuvant therapy arms: use of MAO inhibitor therapy including selegiline or rasagiline within 30 days prior to taking study drug and first posturography evaluation.

## Study Visits

### Screening

Patients will be evaluated by movement disorders expert who will confirm diagnosis of Parkinson disease as stipulated in inclusion criteria. Blood pressure and heart rate, as well as a complete Unified Parkinson Disease Rating Scale (UPDRS) evaluation will be performed.

### Randomization

Subjects will be randomized by facility pharmacy to either placebo or rasagiline 1 mg daily for 8 weeks. There will be 5 rasagiline monotherapy and 5 matching placebo subjects as well as 5 rasagiline adjuvant therapy and 5 matching placebo subjects.

Computerized dynamic posturography evaluation:

Subjects will be evaluated at baseline prior to taking first tablet as well as at 4 weeks and 8 weeks. In the adjuvant therapy arm, subjects will be evaluated on rasagiline but in a functional OFF state for their other PD medications.

Functional OFF will be defined as at least 24 hours post last dose of long acting dopamine agonist or levodopa extended or continuous release formulations, at least 8 hours post last dose of immediate release dopamine agonist, immediate release levodopa, or trihexyphenidyl.

Subjects will undergo computerized dynamic posturography testing via apparatus manufactured by Neurocom, Inc. This device is clinically used for measurement of balance impairment. It measures body sway under varying visual and vestibular cues [3]. Motor control test (MCT) measuring body sway during small translations of the support surface will also be used.

### Conclusion of study

Subjects will stop taking rasagiline or placebo once their 8 week supply runs out.

### Power analysis to determine the number of subjects

To estimate number of subjects needed for the posturography test, we performed a power analysis using the software G*power [4]. For inputs, data from the effect of dopamine depleter tetrabenazine on posturography in Huntington’s disease was used [5].

The strategy composite score here during the tetrabenzine ON state was 79.2, during tetrabenzine OFF state it was 63.5. Averaged SD for these two outcomes was 11.5 and the total sample size was 10. These data were used to calculate the anticipated effect size. This input provided the Effect Size f = 0.6826087 (Table 1).

**Table 1.**
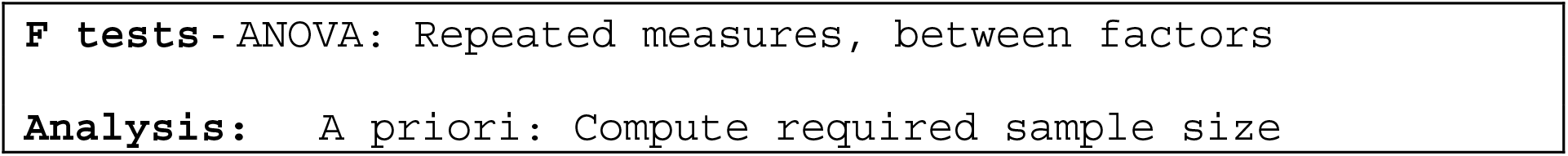
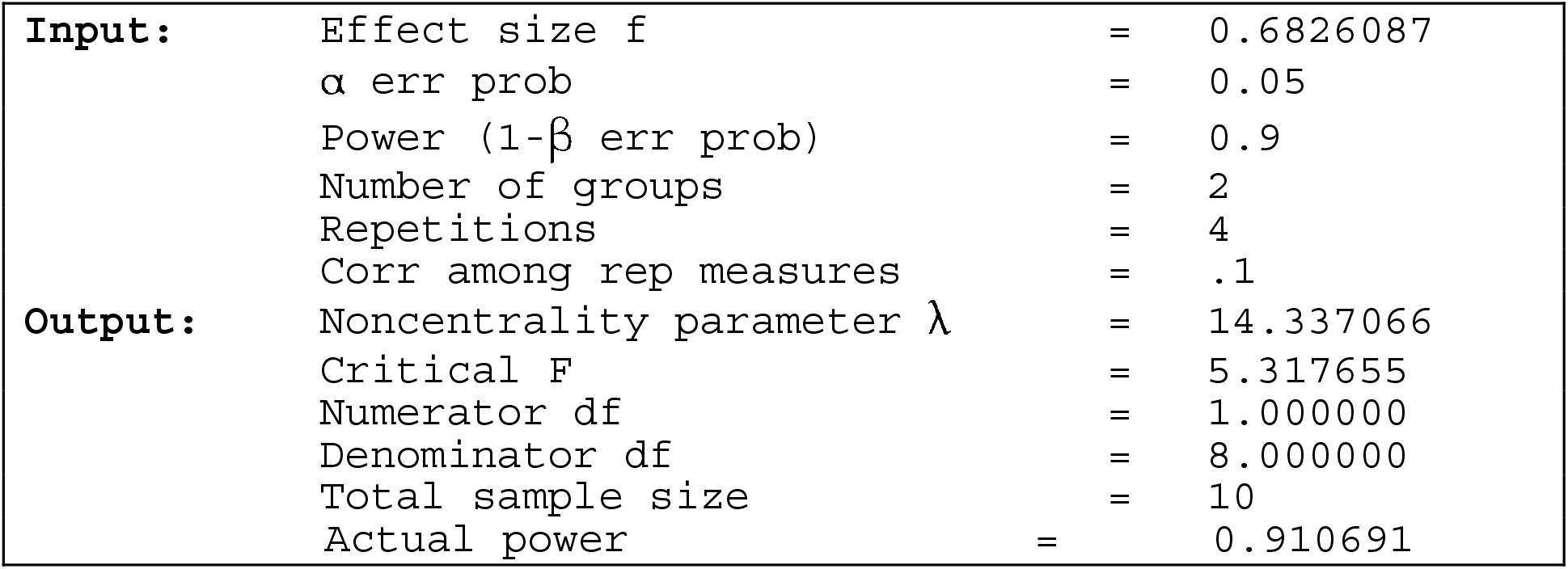
G*power software inputs and outputs for statistical power.

|  |
| --- |
| **F tests** - ANOVA: Repeated measures, between factors |
| **Analysis:** A priori: Compute required sample size |

Table 1. G\*power software inputs and outputs for statistical power
|  |  |  |  |
| --- | --- | --- | --- |
| **Input:** | Effect size f | = | 0.6826087 |
| | $\alpha$ err prob | = | 0.05 |
| | Power (1-β err prob) | = | 0.9 |
|  | Number of groups | = | 2 |
|  | Repetitions | = | 4 |
|  | Corr among rep measures | = | .1 |
| **Output:** | Noncentrality parameter $\lambda$ | = | 14.337066 |
|  | Critical F | = | 5.317655 |
|  | Numerator df | = | 1.000000 |
|  | Denominator df | = | 8.000000 |
|  | Total sample size | = | 10 |
|  | Actual power | = | 0.910691 |

Input of the effect size value into the G*power with requested α=0.05 and power (1 - β)=0.9, weighing in the repeated measurements (before and after rasagiline) with very low correlation among repeated measures resulted in returned total sample size of 10 subjects Table 1 (please see the table of calculated inputs and outputs below). This total number will be randomized into 2 groups of subjects: rasagiline monotherapy and placebo study arm. Similarly, there will be 10 subjects in the study arm with rasagiline as an adjuvant therapy versus placebo.

### General approach for statistical evaluation of the study data

The output of the posturography testing are computer-calculated scores. Though these scores are discrete (do not represent a continuous variable), they still may fit the requirements of parametric statistical tests. Thus, we will first test the raw scores for normal distribution using a Kolmogorov-Smirnov test versus dataset with known Gaussian distribution. If the scores are normally distributed, we will determine whether data variance is approximately equal in all tested groups. If these conditions are met, we will continue data evaluation using repeated measures ANOVA with between factor of drug (levels: rasagiline, placebo) and within factor of treatment (levels: before, after). Level of significance will be preset to 0.05. Since no multiple comparisons are planned, there will be no adjustments of p.

If we do not find the Gaussian distribution of scores or if the variances among the groups are unequal (difference of one order of magnitude or more), we will calculate non-parametric statistics to determine differences. For this purpose we will use Wilcoxon Signed Rank test and evaluate separately rasagiline and placebo groups with p preset to 0.05.

## Data Availability

Given the absence of funding, data gathering could not commence. No data was gathered.

## Abbreviations

COMT: Catechol-O-Methyltransferase
HD: Huntington’s disease
MCT: Motor control test
PD: Parkinson’s disease

## Declarations

### Ethics Approval and Consent to Participate

Clinical trial registered at clinicaltrials.gov as NCT07077187. The study was approved by the New York Medical College Office of Research Administration Institutional Review Board under tracking number L-10,768.

### Consent for Publication

Not applicable

### Author Contributions

RF Conception, writing of manuscript. All authors have reviewed the manuscript.

### Data Availability

Not applicable (this manuscript does not report data generation or analysis).

## Acknowledgements

None

## References

1. Wood BH, Bilclough, Bowron A. Incidence and prediction of falls in Parkinson’s disease: a prospective multidisciplinary study. J Neurol Neurosurg Psychiatry 2002;72:721–725

2. Ebmeier KP, Calder SA, Crawford JR, et al. Mortality and causes of death in idiopathic Parkinson’s disease: results from the Aberdeen whole population study. Scott Med J 1990;35:173–5.

3. Ondo WG, Almaguer M, Cohen H. Computerized posturography balance assessment of patients with bilateral ventralis intermedius nuclei deep brain stimulation. Mov Disord. 2006;21:22243–7.

4. Faul F, Erdfelder E, Lang AG, Buchner A. (G*Power 3: A flexible statistical power analysis program for the social, behavioral, and biomedical sciences. Behavior Research Methods 2007:39(2), 175–191. 10.3758/BF03193146.

5. Fekete, R., Davidson, A., Ondo, W.G., and Cohen, H.S. Effect of tetrabenazine on computerized dynamic posturography in Huntington disease patients. Parkinsonism and Related Disorders 2012;18(7):896–8.

